# Considerations of HIV PrEP Among Heterosexually Active Women and Men: Results from a Qualitative Study in New York City

**DOI:** 10.1101/2025.05.08.25326819

**Authors:** Étienne Meunier, Andrea Ávila, Paul Kobrak

## Abstract

Although 22% of new HIV diagnoses in the United States are attributed to heterosexual contact, uptake of pre-exposure prophylaxis (PrEP) remains low among heterosexually active women and men, and information about PrEP considerations in this population is scarce. We report on a cross-sectional qualitative study that explored attitudes towards PrEP among a diverse sample of 50 heterosexually active cisgender adults (31 women, 19 men) in New York City. We categorized factors influencing PrEP attitudes along social-ecological levels: societal (e.g., social marketing), community (e.g., health care organizations and providers), interpersonal (e.g., friends and sexual partners), and individual (e.g., personal beliefs and preferences). About two-thirds had previously heard of PrEP, but a few had inaccurate or lacking knowledge of it, and none had ever used it. Many participants had learned about PrEP through marketing and social interactions, from which many perceived that PrEP was mainly indicated for gay or bisexual men and transgender individuals. Most participants had never discussed PrEP or received information about it in health-care settings, even though the majority had recently been screened for HIV or sexually transmitted infections. Participants generally felt PrEP was not relevant to them because they perceived themselves at low risk for HIV, thinking PrEP would be indicated for people who have high numbers of sex partners or partners with HIV. Some participants said they would view a potential partner using PrEP positively (e.g., as responsible), while others raised concerns (e.g., about their presumed behaviors). Concerns with side effects and long-term drug toxicity were common, and a few participants expressed anti-medication beliefs. Although awareness seems high, PrEP appears to not have yet entered the repertoire of HIV prevention options for heterosexually active New Yorkers. PrEP promotion among this population could benefit from: messaging targeted to heterosexual adults; ensuring health-care providers inform all sexually active patients about PrEP; and clarifying for practitioners and the public that PrEP is an option for any sexually active person, even those who do not report substantial HIV risk.

## Introduction

In 2022, 22% of new HIV diagnoses in the United States were attributed to heterosexual contact, making it the second largest transmission category after male-to-male sexual contact [1]. Both nationally and in New York City (NYC), heterosexual contact is the main transmission category among women, and new diagnoses disproportionately affect Black and Latina women [2,3].

Pre-exposure prophylaxis (PrEP) is a highly effective HIV prevention method that involves using antiretroviral medicines before exposure [4]. Increasing PrEP coverage has been associated with a decrease in new HIV diagnoses in the United States since FDA approval in 2012 [5]. However, in recent years HIV incidence has only decreased significantly for cases attributed to male-to-male sexual contact [2]. Effectively promoting PrEP use beyond gay, bisexual and other men who have sex with men (MSM) could help achieve similar decreases in HIV incidence among heterosexual women and men.

In the United States, PrEP can be prescribed as an injection every two months, a daily pill, or (for MSM only) pills taken before and after sex. Since 2014, U.S. Centers for Disease Control (CDC) guidelines have recommended providers offer PrEP to sexually active patients who in the last six months a) had a partner with unsuppressed HIV, b) had one or more sex partners of unknown HIV status and did not always use condoms, or c) who had a diagnosis of gonorrhea, syphilis, or (for MSM) chlamydia. To increase PrEP awareness and decrease barriers to discussing it between providers and patients, the CDC’s updated 2021 guidelines added that providers should inform all sexually active adults and adolescents about PrEP, and that “patients who request PrEP should be offered it, even when no specific risk behaviors are elicited” (p. 22) [6].

PrEP awareness and use have been low in groups other than MSM: in the 2019 CDC National HIV Behavioral Surveillance survey, 32% of heterosexually active adults were aware of PrEP and less than 1% had used it [7]. In NYC, the 2016 Community Health Survey found that only 22% of women and 20% of men who only had partners of different gender were aware of PrEP compared to 83% of MSM [8]. Analyses of U.S. prescription data have shown that women who could benefit from PrEP were much less likely to use it than men [9,10]. Though New York State has assistance programs to help residents access PrEP at no cost, uptake has still been much slower among women than men [11].

Prior studies have identified several barriers to PrEP uptake among various populations such as lack of awareness, low perceived HIV risk, concerns about cost and insurance coverage, and worries about side effects [12–14]. However, research on PrEP acceptability has largely focused on MSM, and there is limited knowledge on this topic specific to cisgender heterosexually active adults, particularly heterosexual men [15]. Researchers have also often relied on surveys to ask participants if they endorsed specific barriers or facilitators, and more qualitative research could help understand how people perceive PrEP and their thought process when considering its use.

Although there is a growing body of qualitative research about PrEP among cisgender women [16–25], only two studies we know of included heterosexual men [26–29], and many were among participants who had little or no prior knowledge of PrEP. In these studies, some participants said they had not considered PrEP because they did not know anyone taking it, because they thought it was mostly for MSM, or because they anticipated others might stigmatize them for using it. Such commonly identified barriers may have subsided with continued PrEP promotion and education. As awareness and knowledge of PrEP increases, up-to-date data is important to understand how attitudes are evolving, and which barriers to use might be persisting.

Understanding heterosexually active adults’ attitudes towards PrEP may help inform how public health can address the persistence of heterosexual transmission of HIV. To update and deepen our understanding on this subject, we present qualitative data about PrEP considerations among a sample of heterosexually active adults in NYC.

## Methods

The data we present in this paper were collected by the NYC Health Department as part of a cross-sectional qualitative study aimed at improving HIV and STI prevention efforts for heterosexually active adults in NYC. Between August 2023 and March 2024, we interviewed 50 heterosexually active adults using the NYC Health Panel, a research panel of approximately 35,000 adult New Yorkers assembled by the NYC Health Department [30]. We recruited from among a subset of panel members who had taken a sexual-health survey in the first half of 2023. Based on their survey answers, respondents were eligible for the qualitative study if they: a) were cisgender women or cisgender men; b) resided in NYC; c) reported at least one opposite-gender sexual partner in the prior 12 months; d) reported two or more total sexual partners (of any gender) in the prior 12 months or a sexually transmitted infection (STI) diagnosis in the prior 12 months or had ever used PrEP; e) completed the survey in English or Spanish; and f) agreed to be contacted about future studies.

A total of 308 survey respondents met these inclusion criteria. We aimed to interview 50 participants based on budget constraints and anticipated numbers required to obtain sufficient information on the study topics. We used a purposive sampling strategy to select a sample that was diverse and prioritized demographic groups that have elevated HIV incidence in NYC by including at least 30 women, 18 participants who identified as Black, 12 participants who identified as Latina or Latino, participants from all five NYC boroughs, and at least 12 participants in each of three age groups (18 to 29, 30 to 39, and 40 or older). Within each demographic category, we randomly contacted recruits until we completed 50 interviews.

Three interviewers conducted the interviews (49 in English and 1 in Spanish), which lasted between 28 and 80 minutes (average: 50 minutes). Interviews were audio recorded, transcribed, and (when applicable) translated into English. Participants received a $50 electronic gift card after completing their interviews.

The questionnaire began with a few close-ended questions to collect updated demographic information. Then, using a semi-structured interview guide, participants were asked a series of open-ended questions about their recent sexual behaviors, concerns about sex, HIV/STI preventative behaviors, HIV/STI testing history and habits, and perceptions of biomedical HIV prevention methods (emergency post-exposure prophylaxis [PEP], PrEP, and HIV treatment as prevention). Data presented in this article focus on participants’ responses to the series of questions about PrEP (included as a supplemental file). We first read a brief definition of PrEP and asked participants if they had ever heard of it. If so, we asked what they had heard and whether they had ever discussed PrEP with a health-care provider or anyone else. Regardless of prior awareness, all participants were asked if they would consider using PrEP and how they would feel if a sex partner used it.

In a first round of analysis, the study team organized the transcribed data along the principal domains of the study (e.g., concerns about sexual health, HIV/STI testing, PrEP, etc.). Through an iterative process of preliminary coding and debriefing among team members, the investigators developed a common definition of structural codes that identified all of the text related to each study domain [31]. Each interviewer then applied the structural codes to the interviews they had conducted, and one other team member revised their coding. Discrepancies in coding were resolved through discussion among the investigators. For the analysis presented in this paper, the lead author further examined the excerpts labelled with the “PrEP” structural code. Based on a qualitative content analysis strategy [32,33], the author coded these sections with the goal of producing an exhaustive account of participants’ responses organized into categories that best contained the data. The development and refinement of these categories were periodically discussed among the investigators until they agreed that the results presented below provided an accurate account of the participants’ perspectives on PrEP.

During the analysis process, the authors found that a social-ecological perspective provided a relevant organizing framework for the results. The social-ecological perspective emphasizes that health behaviors are not solely dependent on individuals’ decisions, but happen in interaction with societal forces, cultural context, communities and their environment, and interpersonal dynamics [34]. This perspective has been used to examine attitudes and barriers to PrEP among various populations [13,22,28,35].

## Results

### Participants characteristics

Table 1 shows categorical data for the 50 interviewed participants. Reflecting our sampling strategy, our sample included more women than men, and more participants who identified as Black or Latino than other racial/ethnic groups. More than half of participants resided in NYC areas with the highest rates of HIV diagnoses in 2023^1^. Although most participants had reported two or more sex partners in the year prior to the survey used for recruitment, during our qualitative interviews that took place up to a year later, 22 (44.0%) participants reported only one past-year sex partner. Only one participant reported having been diagnosed with HIV; he was also the only man in the study who identified as bisexual and reported sex with women and men. Among the 49 participants who had never been diagnosed with HIV, the majority (34; 69.4%) had been tested in the prior year and 4 (8.2%) had never been tested or were unsure if they ever had.

**Table 1.**
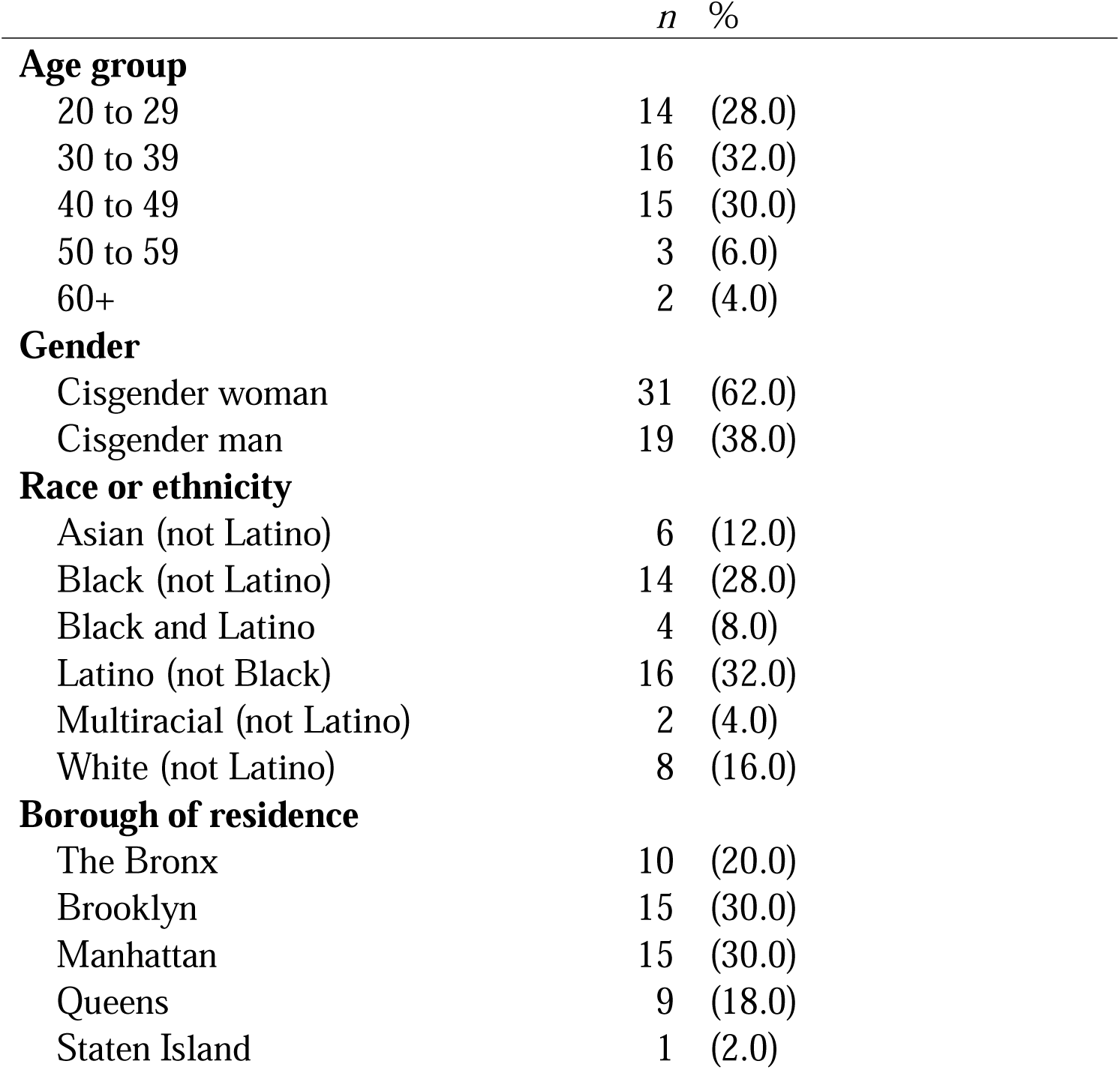
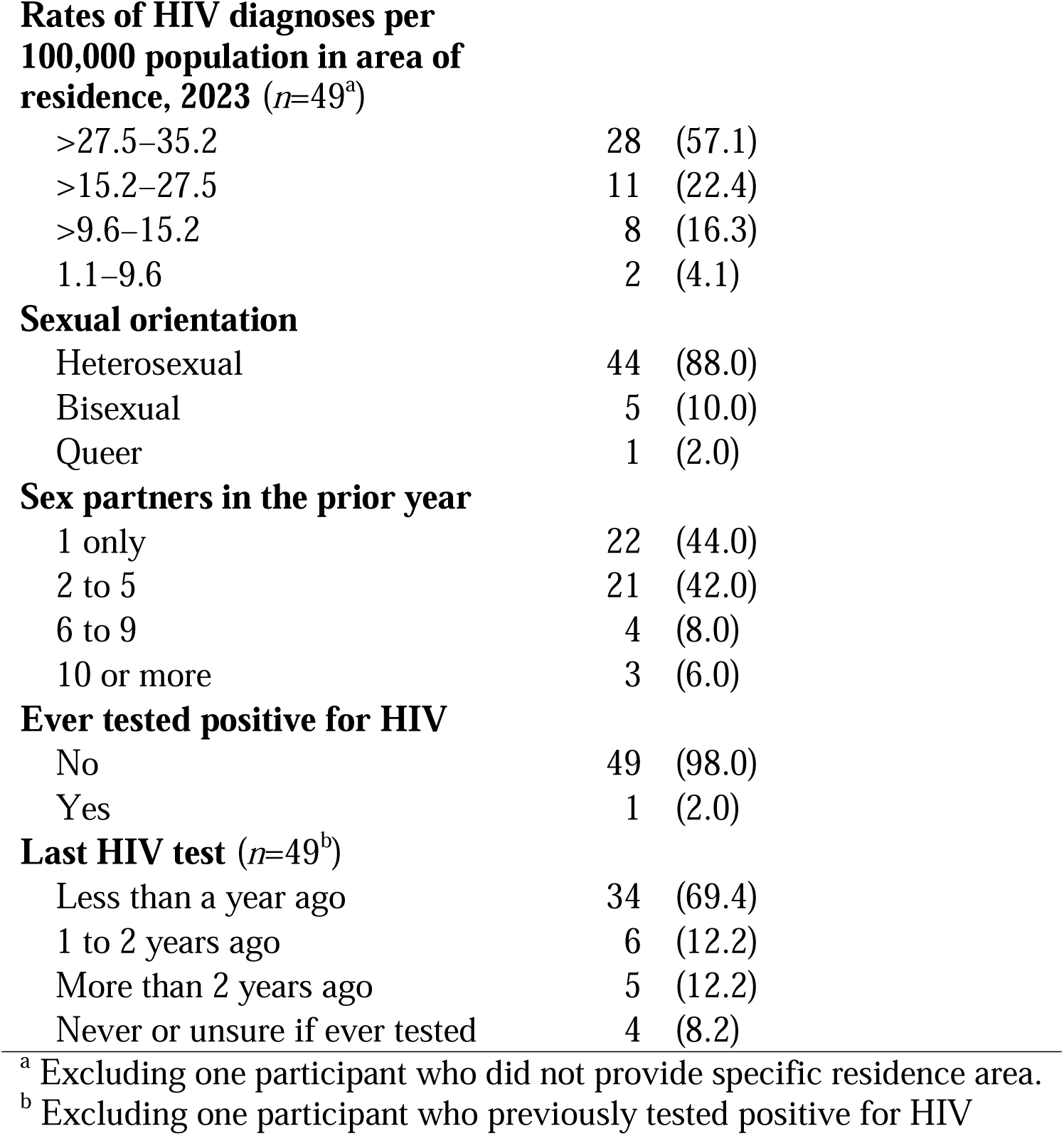
Categorical data from 50 qualitative interviews.

|  | *n* | % |
| --- | --- | --- |
| **Age group** |  |  |
| 20 to 29 | 14 | (28.0) |
| 30 to 39 | 16 | (32.0) |
| 40 to 49 | 15 | (30.0) |
| 50 to 59 | 3 | (6.0) |
| 60+ | 2 | (4.0) |
| **Gender** |  |  |
| Cisgender woman | 31 | (62.0) |
| Cisgender man | 19 | (38.0) |
| **Race or ethnicity** |  |  |
| Asian (not Latino) | 6 | (12.0) |
| Black (not Latino) | 14 | (28.0) |
| Black and Latino | 4 | (8.0) |
| Latino (not Black) | 16 | (32.0) |
| Multiracial (not Latino) | 2 | (4.0) |
| White (not Latino) | 8 | (16.0) |
| **Borough of residence** |  |  |
| The Bronx | 10 | (20.0) |
| Brooklyn | 15 | (30.0) |
| Manhattan | 15 | (30.0) |
| Queens | 9 | (18.0) |
| Staten Island | 1 | (2.0) |

**Rates of HIV diagnoses per 100,000 population in area of residence, 2023 ( $n=49^a$ )**
|  |  |
| --- | --- |
| >27.5–35.2 | 28 (57.1) |
| >15.2–27.5 | 11 (22.4) |
| >9.6–15.2 | 8 (16.3) |
| 1.1–9.6 | 2 (4.1) |

**Sexual orientation**
|  |  |
| --- | --- |
| Heterosexual | 44 (88.0) |
| Bisexual | 5 (10.0) |
| Queer | 1 (2.0) |

**Sex partners in the prior year**
|  |  |
| --- | --- |
| 1 only | 22 (44.0) |
| 2 to 5 | 21 (42.0) |
| 6 to 9 | 4 (8.0) |
| 10 or more | 3 (6.0) |

**Ever tested positive for HIV**
|  |  |
| --- | --- |
| No | 49 (98.0) |
| Yes | 1 (2.0) |

**Last HIV test ( $n=49^b$ )**
|  |  |
| --- | --- |
| Less than a year ago | 34 (69.4) |
| 1 to 2 years ago | 6 (12.2) |
| More than 2 years ago | 5 (12.2) |
| Never or unsure if ever tested | 4 (8.2) |

<sup>a</sup> Excluding one participant who did not provide specific residence area.
<sup>b</sup> Excluding one participant who previously tested positive for HIV

### Socioecological organization of participants’ responses about PrEP

When asked if they had heard about PrEP prior to the interview, most participants (34; 68%) said they had, though some only had a vague idea of what it was, and none had ever used it. All participants were invited to share their thoughts about the strategy whether they had previously heard of it or not. Table 2 provides an exhaustive list of content categories reflecting all participants’ responses about PrEP organized along: a) awareness and knowledge of PrEP, b) beliefs and perceptions of this prevention strategy; and c) considerations for taking it or not. These content categories are further organized along socioecological levels: societal (e.g., media, social marketing, and health-care policies); community (e.g., education, work, and health-care utilization); interpersonal (e.g., social interactions and sexual and romantic partnerships), and individual (e.g., personal attitudes about HIV prevention and medicine).

**Table 2.** Social-ecological organization of responses about PrEP among a sample of 50 heterosexually active women and men in New York City.

|  | **Awareness and knowledge sources** | **Beliefs and perceptions** | **Considerations for use** |
| --- | --- | --- | --- |
| **Societal** | •Social marketing & commercials (17)<br>•Media (5) | •PrEP is for MSM (11) | •Insurance or cost concerns (6) |
| **Community** | •Work or education in health care (5)<br>•Info at health-care organizations (3)<br>•Discussed with health care provider (5) | •Should be more widely promoted (3) | •Anticipated provider barriers (3) |
| **Interpersonal** | •Social discussions about PrEP (16)<br>•People they know using PrEP (11) | *About a potential partner using PrEP:*<br>•Would want to know why (17)<br>•Might have partners with/at risk for HIV (8)<br>•Might have HIV (5)<br>•Might have concurrent partners (5)<br>•Might be MSMW (3)<br>•Worry about efficacy or their adherence (4)<br>•Is responsible about sexual health (16) | •Not needed in committed relationships (11)<br>•Relevant if multiple partners (9)<br>•Relevant if partner with HIV (17) |
| **Individual** | •Would need to learn more about PrEP (7) | •Medication hesitancy or skepticism (7)<br>•Concerns about efficacy (2)<br>•Misunderstandings about PrEP (8) | •Side effects concerns (14)<br>•Medication interaction or overload (2)<br>•Self-efficacy concerns (4)<br>•Low perceived HIV risk (11)<br>•Prefer relying on condoms (7)<br>•Some willingness to use it (9) |

Notes: Numbers in parentheses indicate how many participants expressed each point.
MSM: men who have sex with men; MSMW: men who have sex with men and women.

### Societal factors: public messaging on PrEP

Many participants mentioned seeing *social marketing and commercials*^2^ about PrEP including ads in NYC public transportation: “You walk down the street and you’ll see an ad for PrEP, or you’ll be on the subway and you’ll see something about testing. I feel like we’re sort of inundated with messaging in New York” (Laura^3^: 40s, white). TV commercials from pharmaceutical companies also fostered basic awareness of PrEP: “I’ve seen ads on TV, like that you could take them [medications] to prevent [HIV]” (Ivan: 20s, Latino). Some had heard about PrEP through various *media* including social media, television shows, and journalism; “I’ve seen it on the news” (Samuel: 20s, Latino).

However, many participants admitted not paying much attention to messaging about PrEP. Alfred recollected hearing about PrEP but felt the information was not relevant to him: “I don’t think it concerns me because I wouldn’t take any kind of medication to take chances with somebody” (Alfred: 60s, Latino). Participants felt they were not the target audience of PrEP messaging and many believed that *PrEP is for MSM*: “Most of the advertising is geared towards gay men” (Nicole: 40s, Latina). Consequently, most participants never felt like learning more about PrEP after seeing messaging about it:

> I’ve seen advertisements around the city, from which I have always understood that PrEP has been used predominantly by members of the LGBTQ community. … I’ve always seen advertisements that seem to be targeting particularly that community, so I’ve never really been interested in exploring any of that. (Omar: 20s, Black)

### Community factors: Health care

Several participants mentioned learning about PrEP through their *work or education in health care*: “I actually became a [medical professional] a couple of years ago, and we were made aware of PrEP and PEP” (Andre: 30s, Black and Latino). However, only a few participants said they had received information about PrEP during health-care visits. Some had seen or received *informational materials at health-care organizations*: “I was at a city-run clinic getting tested. They had a bunch of flyers and brochures and a TV in the waiting room. That’s where I first saw it [PrEP]” (Jackie: 20s, Latina).

Only five participants said they had *discussed PrEP with a health care provider*. Two of them said they had initiated this discussion. Henry, who reported 15 different sex partners in the prior year, said he felt curious about PrEP after learning about it on a subway ad. He asked his provider about it, but was left to do his own research:

> I did ask the doctor about it. … I had first learned about it in the New York City subway and the idea was fascinating and revolutionary to me, so I wanted to know the scientific mechanism, how it worked. … The doctor did not really seem to know anything about it. … I satisfied my curiosity to a degree through the internet, and then it kind of faded for me because it never became a possibility of “I have to use this” … because I don’t have any sexual partner who is HIV positive. (Henry: 40s, Asian)

Only three participants—all of them Black women—said their providers had initiated a discussion about PrEP. They described their providers informing them about PrEP without explicitly suggesting they use it: “They weren’t recommending it to me. They were just discussing” (Michelle: 40s, Black and Latina). Brandy thought that being a Black woman explained why two different providers told her about PrEP. Yet, like others who had been informed, she appreciated knowing she could access it:

> Homosexual men are more eligible; resources are more available for them than it is for women. But, as a Black woman, as an African American woman, I’ve seen those resources being sent to me too. … They offered the information: “Have you heard of it? This is what it’s for. If you have any concerns or questions about it, please let us know.” Then they give you a pamphlet, but no long discussion. … It has happened with two providers. … I appreciated being informed and I also appreciated the fact that they made it known that that was something that I could look into. Knowing that there’s a resource that I have access to is always mentally better. (Brandy: 30s, Black)

Some participants *anticipated provider barriers* to PrEP use, like Joanna who had heard that it could be difficult to get a provider to prescribe PrEP:

> If you only have a couple of partners here and there, the doctors will say, “Oh, it’s for high-risk patients, those who are very active.” I’ve had friends in the gay community that had to lie and up their amount of engagement and their risk just to be able to get PrEP. I’m not active all the time, just moments, and I don’t know that I would qualify for PrEP. (Joanna: 40s, multiracial)

Only one participant expressed a clear willingness to start using PrEP. Eve had recently been diagnosed with an STI, which she felt she might have acquired from an ongoing male partner who told her he also had sex with men and who refused to use condoms with her. However, because her providers had refused to vaccinate her against human papillomavirus (HPV) based on her age, she was convinced they would not want to prescribe her PrEP:

> They keep telling me, “You don’t need it [HPV vaccine].” And I’m like, “Yes, I do. I know my history and I know my behavior, and I’m telling you that this is something that would benefit me,” and they’re like, “Nope, it’s not.” … There are assumptions that are getting made that, by this time, I would be married, the two of us would only be having sex with each other. But I’m telling you that’s not my life. … I’d take PrEP today if I had the opportunity, … because I know my own history and I know my own activities. … She [my provider] would be [reluctant] because if she won’t give me Gardasil— (Eve: 40s, Black)

Some participants also anticipated barriers related to health-care policies (a societal factor) and expressed *insurance or cost concerns*. They anticipated it might be difficult to get the medication covered by their health-care insurance: “If it wasn’t covered by my insurance, I wouldn’t take it. Or if it wasn’t free” (Natalie: 40-year-old white woman). Indeed, some participants had heard from others that insurers could make PrEP difficult to access: “My friend was sharing his frustration because he was supposed to get PrEP at some point recently, but there was something with his insurance that prevented it from happening” (Camille: 20s, Black and Latina).

### Interpersonal factors: social interactions and partners

#### Social discussions about PrEP

Several participants described *social discussions about PrEP* though which they learned more about it or, in some cases, informed others. For example, Maya talked about PrEP to a friend: “My Korean friend, her brother just came out as gay. Korean society is very conservative. There’s a lot of homophobia. … So, I was telling [her] about PrEP and some of the things that he may have access to” (Maya: 30s, Asian).

Some of these discussions were with *people they knew who were using PrEP*. In most cases, these were gay men or transgender persons (only one knew a cisgender woman using PrEP), which further reinforced participants’ belief that PrEP was not relevant to them: “I have male gay friends who take it. I think it’s rarely suggested to people who identify as female. I don’t think I’ve ever really heard of any of my girlfriends taking it” (Katrina: 20s, Asian).

#### Sexual and romantic partners

Many participants determined PrEP was not relevant to them based on their current sex life and partners. Some of them said PrEP was *not needed in committed relationships*: “Based on my current relationship, I don’t think that would need to be the case. … Not my current situation” (Joe: 40s, white). In the survey used for recruitment, most participants had reported two or more past-year sex partners, but during the interview many said they were now in committed relationships and had not had other recent sex partners. Some said they might consider PrEP if they were to start seeing new partners again:

> If I start seeing a new person. If my partner and I break up, we no longer stay together and then I’m seeing a new person. Or if I decide I want to be non-committal and date: be, I guess, polyamorous, be in poly relationships. I think that’s when I would concern myself with it a bit more. (Brandy: 30s, Black)

In a similar vein, many participants felt like PrEP was mostly *relevant for people with multiple sex partners* or who have frequent condomless sex:

> It sounds good for someone who’s sleeping with multiple people and having a lot of unprotected sex. Then I guess that would be a good thing, PrEP; it could prevent you from getting HIV. I think that that could be good, but I don’t think it’s necessary if you’re not having multiple partners and things like that. (Michelle: 40s, Black and Latina)

Many participants also thought that PrEP might be *relevant if they had partners with HIV*: “I would say if I met someone that was telling me that they’re HIV positive, then I think that would be something I would take into consideration” (Amarylis: 30s, Latina). However, some participants admitted they might still be concerned about having partners with HIV: “I think [I might be interested in PrEP] if I met someone who happened to be HIV positive and I somehow got past my own fear of contracting it myself” (Pete: 50s, Black).

Two participants (other than the one who had HIV) said they ever had sex with a partner they knew had HIV. Both of them did so after PrEP had become available, but neither used it because they relied on the fact that their partners were virally suppressed and could not transmit HIV through sex. However, both said they would consider PrEP if they were to have a partner with HIV again: “If I were to date somebody that was HIV positive again, which I absolutely would, I would consider taking PrEP” (Laura: 40s, white).

#### Perceptions of potential partners who use PrEP

Only the participant who had HIV reported ever having a sex partner that he knew to be using PrEP. When asked how they would feel if a sex partner revealed they were using PrEP, participants expressed both concerns and reassurance in roughly equal measure. A common reaction was that they *would want to know why* that person was using PrEP. Even though they understood that PrEP prevents HIV acquisition, some participants felt that a partner using it might present higher risks:

> I would maybe be questioning why they feel like that’s the thing that they should be doing. I’d probably be a bit more cautious towards having unprotected sex with them. I know it’s irrational, but it’s like a stigma about their behavior. I know it’s counterintuitive in many ways because it means that there is probably less chance of them having it and passing it on to you. That’s the way I would think about it, but it’s counterintuitively irrational. (Luis: 40s, Latino)

In many cases, participants presumed these partners would be using PrEP because they *might have partners with or at risk for HIV*: “HIV is something that’s present in their life and they’re either concerned with transmitting it or acquiring it. Something to be careful with” (Sam: 20s, Latino). Some participants were specifically concerned that people using PrEP *might have HIV*: “If they told me they were taking PrEP, I would definitely be quite a bit concerned. I know that doesn’t sound intelligent because it’s for preventing, but it does spark the theory of “Oh, does this person have HIV?” (Troy: 20s, Black).

Other concerns included that a partner using PrEP *might have concurrent partners*: “I would think that they were very promiscuous if they told me that, even though it might be stigmatizing or stereotyping. To be honest, that’s what I’ll think” (Monique: 40s, Black). Based on such presumptions, some participants said they might avoid a partner using PrEP: “That means that he’s a liability, that he’s out there having sex with multiple women. … That’s not appealing. That would be someone that I would not want to have sex with, because he has risky sexual behaviors” (Jenny: 40s, Black and Latina).

Some women said they would think that partners using PrEP *might be men who have sex with both men and women*. Although they acknowledged taking PrEP would be responsible, they had lingering concerns about a partner possibly having sex with men:

> I would assume he has male and female partners. … There’s that perception of gay sex potentially spreading HIV—obviously not necessarily true, but I think that perception creates a bit of concern. It’s kind of funny that that person being more responsible could elicit a “That kind of concerns me” response. That’s a little sad … but I think my initial reaction might be like, “Should I be worried?” (Katrina: 20s, Asian)

Talking about potential partners using PrEP, some participants brought up *worries about efficacy or adherence*: “What if they forget their pills? What if they just stop taking it and don’t say anything?” (Arielle: 30s, Latina). Presuming people using PrEP had other partners with HIV, efficacy could be a concern: “I would probably assume that they had been sleeping with someone who had HIV. That part would concern me because then I’m like, ‘Does it actually work? Is it actually safe?’ And it would probably make me nervous” (David: 20s, white).

Although many participants expressed concerns, a common positive reaction toward PrEP users was that they are *responsible about sexual health*: “I would think that they were making a smart decision, almost like if I was speaking with another person who was taking birth control” (James: 30s, Black). Some participants thought a partner on PrEP would be preferrable to others: “Honestly, that’s a green flag because you know that they’re being proactive about their sexual health and most likely getting tested regularly as well. You can’t say the same for any random person” (Camille: 20s, Black and Latina).

### Individual factors: medication beliefs, perceived risk, and knowledge gaps

#### Concerns with medication

At the individual level, participants had various thoughts about medications that informed their considerations for using PrEP. Commonly cited were *side-effects concerns* and worries about long-term drug toxicity: “I would want to know a little bit more about potential side effects of putting something in my body every single day for a while” (Lila: 20s, Asian). Other participants were more specifically concerned with *medication interactions or overload*: “I don’t think it’s good to be taking another medication because, for me, I have high blood pressure, I have diabetes. I’m not going to put another pill on top of the pills if I don’t need it” (Michelle: 40s, Black and Latina).

A few participants expressed *medication hesitancy or skepticism*. Some said they were generally reluctant to use medications of any kind unless they had to: “I’m trying not to take as much medication unless I have a health threat. … I don’t like taking medication” (Brianna: 30s, Black). Such participants preferred what they considered more holistic ways of maintaining health:

> I’m not interested really in taking medications, period. … I just don’t like putting, not artificial, but extra things in my body. I like to think my body is kind of tuned on its own. … I prefer to find a natural way versus a manufactured way to deal with an issue or something. … I’m a little more holistic, especially when it comes to medications and treatments than relying on traditional Western medicines. (George: 50s, Black)

As others did when thinking about a hypothetical partner using PrEP, some participants expressed *concerns about PrEP’s efficacy*. The small chances of it failing could be concerning:

> I believe in the magic of statistics. If it’s .001% chance multiplied by .001% chance, there’s still a chance. Yes, I would be concerned about it. Not in an overt way, but it would be in the back of my mind. (Henry: 40s, Asian)

Some participants expressed *self-efficacy concerns*, questioning their own ability to adhere to the medication and keep up with frequent medical appointments: “Maintaining, getting it, and just having to do it on a regular basis means extra work” (Nicole: 40s, Latina). These participants thought taking PrEP would require some effort and felt they might not be up to it:

> I feel like a lot of times it’s like a lot of hassle. And then what if I don’t take it regularly? What if I don’t do what I’m supposed to do? Then that seems kind of bad. … Honestly, just to consistently take it, making sure you don’t, mess up, miss a dose, or misuse. (Brandy: 30s, Black)

#### Risk assessments and prevention preferences

Some participants said they had not considered PrEP because they were in a demographic group at *low risk for HIV*:

> My understanding was that HIV is mostly prevalent in homosexual men and—this is going to sound terrible but—like in Africa or somewhere else that’s not in the US. … I just haven’t even thought to look into HIV prevention or more information about PrEP because I just don’t think of myself as high risk. (Kyra: 20s, Latina)

In some cases, participants balanced their self-assessments of HIV risk against others concerns such as side effects or costs of using PrEP:

> I guess it’s a cost–benefit thing. What’s the cost of it? It’s not something I really see myself needing to pay for because it doesn’t seem like I’m in a specifically risky population or even in a specifically risky behavioral pattern. (Luis: 40s, Latino)

When asked if they considered PrEP, some participants said they did not because they *preferred relying on condoms*: “Not really, because I always like to take precautions” (Alex: 20s, Latino). An advantage of condoms over PrEP was that they also prevent pregnancy and transmission of other STIs:

> I think that at this point in my life, I couldn’t be with someone without taking care of myself with a condom. No matter how much I take that medication, I couldn’t. It’s very ingrained inside my head. … The other person may not have HIV but could have other types of illnesses. I think that a condom would protect me more in that sense, and also for pregnancy prevention. (Matias: 40s, Latino)

Although most participants felt like PrEP would not be relevant to them, a few participants—all of them women—expressed *some willingness to use PrEP*, albeit hypothetically. They acknowledged that PrEP could alleviate worries about HIV, for instance in case of condom failure: “If anything happens, like a condom breaks or a condom is halfway being used. In these kinds of situations, it would be great, but they don’t happen so often” (Joanna: 40s, multiracial). Others remarked that PrEP could be useful during times of heightened sexual activity:

> I wouldn’t mind doing that if I was sexually active with more than one person at a time, or I don’t have a boyfriend and I’m just having sexual activities with random people. … It’s just an extra measure to protect yourself—not another person, yourself. I just feel like that’s actually a good thing. (Vicky: 30s, Latina)

#### Addressing knowledge gaps and improving promotion

Though many participants were aware of it, some displayed *misunderstandings about PrEP*. As mentioned above, some thought that people who use it might have HIV. Others mistook PrEP for the notion that HIV treatment prevents people with HIV from transmitting the virus (also known as *Undetectable equals Untransmittable*). Some participants acknowledged that they *would need to learn more about PrEP* to determine whether they would be willing to use it or not: “I’d be open to learning more about it so that I’m educated on it. … Seeing what’s in it, and if it’s going to be the best thing for me to utilize, if needed” (Julia: 30s, Black).

Tying this individual lack of knowledge to more societal and community factors, some participants said that PrEP *should be more widely promoted* in health care and education. Even if they were not interested in using PrEP, some participants said that they wanted to know about it so they could inform others: “Not for myself, but for my grandkids that are growing. … They don’t teach kids about sex and stuff in schools, so it starts at home” (Hazel: 60s, Black). Another participant expressed disappointment about not having heard of PrEP before: “It should be advertised in doctors’ offices or hospitals, even schools. … It makes me sad that it’s not really advertised. I’m [40-something] years old and it’s the first time I’m ever hearing of any of these” (Sara: 40s, Latina). Finally, Eve emphasized the role of health care providers in disseminating knowledge about PrEP:

> Advertise to women. Tell providers, “Some women are having a far more adventurous life than you anticipate.” We are having sex with men who will not disclose to us who they’re having sex with and we’re not entirely always aware of what’s going on, even if we’re in committed relationships. (Eve: 40s, Black)

## Discussion

Our study among a sample of 50 heterosexually active New Yorkers described how they learned about PrEP, their beliefs and perceptions about this HIV prevention strategy, and their considerations for using it or not. Most participants had some prior knowledge about PrEP but did not perceive it as something that was relevant to them. As HIV incidence attributable to heterosexual contact in the United States has remained fairly stable since PrEP became available, it is important to find ways to increase interest in PrEP among heterosexual adults. As outlined below, our qualitative findings suggest ways to improve PrEP promotion among this population.

We found that PrEP has entered the social environment of heterosexually active adults in NYC though it has not widely become part of their repertoire of HIV prevention strategies. Our reliance on a small purposive sample does not allow us to conclude whether awareness is increasing, but over two-thirds of participants had at least some prior awareness of PrEP, a proportion higher than in prior surveys with heterosexually active adults [7,8]. Participants described hearing about PrEP through ads in public (often in the subway), in television commercials, and in the media, suggesting that promotion efforts have broadened awareness. Many also mentioned friends, family members, or acquaintances they knew were taking PrEP. However, as in prior qualitative studies with heterosexually active women and men [16–18,20,22,27,28], many participants believed PrEP was only relevant for MSM and other LGBTQ individuals, and that more should be done to promote the strategy among heterosexual women and men.

Health-care providers and organizations have an important role in disseminating information about PrEP beyond LGBTQ groups. Since 2021, the CDC recommends that providers inform all their sexually active patients about PrEP [6], but most of the heterosexual women and men we interviewed had not heard about PrEP during health-care visits. Although most of our participants had been screened for HIV and STIs in the last year, few had ever discussed PrEP with a provider. Only three participants, all Black women, had been informed about PrEP at medical appointments. Black women are a priority population for HIV prevention but should not be the only ones to receive information about PrEP during health-care visits. These participants appreciated being informed, which should encourage providers to discuss PrEP with more patients. Women in other studies have also expressed disappointment when realizing their providers should have taken the time to inform them about it [19,28], a sentiment echoed by some of our participants. Further, a few participants in our study thought their providers might not agree to prescribe them PrEP, a barrier that could be alleviated if providers introduced the topic during visits.

Few participants in our study expressed interest in using PrEP. Some women mentioned that it could be helpful during condomless sex, which sometimes happened by choice, by accident, or when a man refused to use condoms. In prior qualitative studies, women similarly expressed that PrEP could be a useful prevention strategy that they could use independently of their male partners [19,36,37]. However, while many women acknowledge the benefits of taking PrEP, these perceptions may rarely translate into uptake. Many participants said they would consider using PrEP if they had a partner with HIV, though some of them also expressed unwillingness to have such partners. Further, the few participants who actually had partners with HIV had chosen not to use PrEP after learning that there was effectively no risk of transmission if their partners were virally suppressed, as was observed in another study with heterosexual women and men [38]. Our in-depth qualitative interviews thus show that even though people can name circumstances that could lead them to consider PrEP, there seem to be other factors preventing them from actually using it when the time comes.

Low perceived HIV risk is one of the most commonly mentioned barriers to PrEP use in the literature [12,14]. Most participants in our study felt that PrEP was not relevant for them because they assessed their risk for HIV to be low. Although people may not always recognize their risk for HIV, we acknowledge that some of our participants could be correct in their self-assessments. Such participants may not have underestimated their risk for HIV, but rather overestimated the requirements for PrEP candidacy, thinking it would only be available to people who have very high numbers of sex partners, who frequently engage in condomless sex, or who have partners with or at high risk for HIV. CDC guidelines [6] do not impose such strict requirements. They suggest that providers offer PrEP to patients who had at least one instance of condomless sex with a partner of unknown status in the past six months, and the exact number of partners does not need to be considered. Providers can prescribe PrEP even when no specific risk behaviors are elicited. However, the initial FDA approval of PrEP indicated it only for adults at high risk for HIV [39], a position that might seems to be enduring among providers and the public. Promoting PrEP use among a broader population may not require increasing people’s perceived level of HIV risk, but clarifying that PrEP can be used by anyone who feels they could benefit from it, even if they do not report substantial HIV risk.

The perception that PrEP comes with important costs—financial and to one’s health— supports the idea that it should only be used in cases of very high need. Our results showed that concerns about side effects and medication toxicity remain a considerable barrier to PrEP uptake, consistent with several prior qualitative studies [16,21,22,24,27,28,36]. Some participants said they generally tried to avoid any type of medications in favor of what they considered more holistic ways to maintain health. Others expressed concerns about the side effects of PrEP or about using too many medications. Overall, participants conveyed a belief that taking fewer medications is preferrable to taking more. PrEP promotion should emphasize what we know about drug safety and how long-term toxicity can be prevented by regular monitoring. Further, more widespread uptake of PrEP may require addressing general social attitudes against medications. Developing modalities of PrEP that do not require a daily pill or frequent injections could also alleviate concerns about the perceived effort required for adhering to the medication [40].

Participants’ perceptions that PrEP use is only justifiable for people at very high risk for HIV also informed how they viewed potential partners using PrEP. Prior studies found that a barrier to PrEP was a worry that others would perceive the person using it as prone to infidelity or promiscuity, or as someone who actually has HIV [16,22–25,27,28]. In our study, some participants expressed such stigmatizing views when asked how they would feel about a partner using PrEP. Based on their own conceptualization of PrEP candidacy, they presumed such partners would be using PrEP because they have multiple concurrent partners or have partners with HIV. Although some participants were concerned about having partners who use PrEP, others viewed them in more positive light, as people who are responsible and careful about their sexual health. Increasing PrEP acceptability could thus also require highlighting positive views of users as people who are diligent about sexual health, regardless of their level of HIV risk.

A few limitations should be considered when interpreting the results from this study. As participants discussed their HIV and STI prevention practices with staff of their local health department, social desirability bias could have influenced them to provide more positive views on PrEP than they would in other contexts. However, since participants expressed a range of positive and negative thoughts about PrEP, we believe this bias might have been minimal. Further, because the interviews were done over the phone (rather than face-to-face) and with the assurance of confidentiality, we believe we created a context in which participants could freely express their opinions. Because of the recruitment and sampling strategy, data from this study cannot be generalized to the heterosexually active adult population. Although the NYC Health Panel aims to recruit a sample representative of the city’s population, enrollment bias could still favor certain groups of people. For instance, several participants mentioned working in health or medical fields, which suggests that people with an interest in health research are more likely to volunteer to be part of a panel and to respond to study invitations. In a more representative sample of New Yorkers, awareness and acceptability of PrEP might have been lower.

Despite these limitations, our study provides updated data about PrEP awareness, knowledge, beliefs, and considerations among heterosexually active women and men in NYC. Drawing from a social-ecological approach, we described how individual attitudes toward PrEP are informed by factors at multiple levels, such as social marketing, health care engagement, social interactions with friends and family, relationships, personal beliefs about medicine, and individual risk perceptions and prevention preferences. Increasing acceptability of PrEP among heterosexually active adults will require efforts at multiple levels, including: social marketing that presents PrEP as a strategy not exclusively for MSM or other LGBTQ individuals, addressing ongoing worries about side effects and long-term drug toxicity, and clarifying that PrEP is not prescribed only to people with specific levels of HIV risk. Providers should be aware of current CDC guidance that recommends they inform all their sexually active patients about PrEP, and future studies could explore provider barriers to discussing PrEP and how to address them.

## Statements and Declarations

## Ethics approval

The study presented in this article was approved by the Institutional Review Board at the New York City Department of Health and Mental Hygiene (protocol #23-051; PIs: Kobrak/Meunier).

## Consent

All individuals provided informed consent prior to participating in the study presented in this article.

## Data availability

Data from this study are not publicly available. Please contact the authors with specific inquiries.

## Authors’ contribution

All authors contributed to the study conception and design, material preparation, data collection, and analysis. The first draft of the manuscript was written by Étienne Meunier and all authors commented on previous versions of the manuscript. All authors read and approved the final manuscript.

## Acknowledgements

The authors want to acknowledge the contributions from colleagues at the NYC Department of Health and Mental Hygiene, including the NYC Health Panel team, the investigators of the Sexual Health Survey in the Time of COVID and MPV, and internal reviewers.

## Supplementary information

### Interview questions related to PrEP

*Considerations of HIV PrEP Among Heterosexually Active Women and Men: Results from a Qualitative Study in New York City*

There is medicine that you take before you have sex that prevents HIV. It is called PrEP, or pre-exposure prophylaxis, and is available as a daily pill or as an injection. If you take PrEP, you have a very low chance of getting HIV even if you have sex without condoms with someone who has HIV.

**Before today, had you heard of PrEP?**

*(If no, never heard of PrEP)*

**Do you think PrEP is something you would want to learn more about?**

Why? Why not?^4^

*(If yes, knew about PrEP)*

**What have you heard about it?**

**Have you ever taken PrEP?**

*(If yes)* What has your experience been like?

**Have you ever discussed PrEP with a doctor or nurse?**

(*If yes)* How did the conversation go?

*(If no)* Would you want to discuss PrEP with a doctor?

**Have you ever discussed PrEP with anyone else, including friends or acquaintances?**

*(If yes)* How did the conversation go?

**Do you personally know anyone who has taken PrEP to prevent HIV?**

(*If yes)* Have you ever talked to them about PrEP? How did the conversation go?

*(All participants)*

**Do you think there could be a situation in which you might want to take PrEP?**

*(If not)* Is there anything that would make you more interested in taking PrEP?

*(If yes or maybe)* Under which circumstances would you consider taking PrEP?

What do you think would be the benefits of taking PrEP?

Do you think there would be anything you would not like about taking PrEP?

**What would you think if a sex partner or potential sex partner told you they took PrEP?**

Would you feel protected because they take PrEP? Would you be concerned?

## Footnotes

1 Information obtained by cross-checking participants’ self-reported zip code of residence with HIV surveillance data [3].

2 Italicized terms refer to the content categories also listed on Table 2.

3 Participant names are pseudonyms assigned by the study team.

4 Indented questions are optional follow-up questions.

